# Health of first- and second-generation immigrant children in England and Wales: serial, cross-sectional study using linked Census and birth registration data

**DOI:** 10.1101/2025.02.18.25322449

**Authors:** Pia Hardelid, Senhao Yu, Jemima Stockton, Rebecca Langella, Kate Lewis

## Abstract

**Introduction:** There is limited research on the health outcomes of children who have migrated, or whose parents have migrated, to the UK.

**Methods:** We used linked birth registration and Census data from the 1991, 2001 and 2011 waves of the Longitudinal Study, covering 1% of the England and Wales population. We derived the prevalence of self-or-caregiver reported limiting long-term illness (2001, 2011 Censuses) and poor health (1991, 2001, 2011 Censuses) among household members <18 years old. We defined immigration background using child and parent country of birth. Logistic regression models assessed the association between immigration background and health outcomes.

**Results:** The percentage of children who were first-or second-generation immigrants increased from 16.6% (20120/121109) in 1991 to 24.7% (28965/117116) in 2011. We found no evidence of an association between immigration background and health outcomes in 1991. In 2011, children born outside the UK were less likely to report a limiting long-term illness (odds ratio 0.67; 95% confidence interval 0.58, 0.76) compared to average. Also in 2011, children born in the UK to UK-born parents or non-UK-born fathers/UK-born mothers had higher odds of reporting long-term illness compared to average (1.25; 1.17, 1.32 and 1.21; 1.08, 1.36, respectively). In both 2001 and 2011, children born in the UK to non-UK-born parents had the highest odds of reporting poor health.

**Conclusion:** Whilst immigrant children with a first-generation immigrant background are less likely to live with limiting long-term illness, evidence-based National Health Service guidelines for supporting second-generation immigrant children are needed.

**What is already known on this topic:**

- Studies of adults have identified a ‘healthy migrant effect’, where immigrants have lower mortality rates than the host population.
- It is not clear if this applies to children or to non-mortality outcomes.

**What this study adds:**

- There was no association between children’s immigration background and prevalence of long-term illness before 2011, or poor health before 2001.
- First generation immigrant children had lower prevalence of limiting long-term illness, and similar prevalence of poor health to non-immigrant children.
- Children born in the UK to UK-born parents or second-generation immigrant children via the father had higher prevalence of long-term illness, and UK-born children whose parents were both born abroad had higher prevalence of poor health.

**How this study might affect research, practice or policy:**

- The National Health Service should develop evidence-based guidelines for how health services can support second generation immigrant children.

## Introduction

European countries require international migration to sustain their economic prosperity and population size.^1^ Almost two in five children born in England and Wales in 2022 had at least one parent born outside the UK,^2^ and 6.6% of children resident in England in the 2021 Census were born outside the UK.^3^ Despite this, international migration history as a social determinant of health has been severely under-researched,^4^ particularly among children. Immigrants in high income countries appear to have lower mortality than the host population^5^ (the ‘healthy migrant effect’ effect^6^). However, it is not clear whether the healthy migrant effect is also apparent in children or for other health outcomes. A systematic review of health service use among first- and second-generation immigrant children living in primarily high-income countries indicated that children were less likely to use healthcare services apart from emergency and hospital services.^7^ This does not appear to be the case in England however, where young children whose parent(s) were born abroad were less likely to be admitted to hospital in an emergency than those with UK-born parents.^8^

There are few UK-based studies of self-reported health among children who have migrated to the UK (hereafter called “first generation immigrants”), or whose parents have migrated to the UK (“second generation immigrants”), with recent studies primarily focusing on adult immigrants.^9-11^ A systematic review^12^ showed that the majority of UK studies on self-reported health, health behaviours or use/experience of health services focused on highly vulnerable groups including unaccompanied asylum-seeking children.

Migration policy in the UK has become increasingly restrictive during the 20^th^ and 21^st^ centuries,^13^ culminating in the UK government’s ‘hostile environment’ policy, first raised during the Conservative/Liberal Democrat coalition government in 2012.^14 15^ Such policies may affect both first- and second-generation immigrant children. More restrictive policies may mean fewer immigrants with pre-existing health problems settle in the UK. However, a more restrictive migration policy, reduced health service access, and the political rhetoric associated with such policies,^16^ may increase discrimination and subsequently worsen the health of immigrants and their children.^17^ For example, mandatory surcharging for secondary care services for non-ordinary residents, including undocumented migrants, was introduced in 2015.^18^

The UN Declaration of the Rights of the Child states that all children have the right to the best possible health.^19^ Everyone in England has a right to register with the National Health Service (NHS) and access primary care and certain other NHS services for free; secondary care is available without charge for individuals ordinarily resident with a legal basis for remaining in the UK.^20^ Evidence on the health problems experienced by first or second generation immigrant children is needed to inform appropriate service provision by health services, schools and other institutions.

We used serial analyses of linked Census and birth registration data from England and Wales to establish the health profile of first- and second-generation international immigrant children between 1991 and 2011, compared to children who are born in the UK with UK-born parents. Our objectives were to describe how the prevalence of caregiver-or self-reported long-term health problems varies between first, second and non-international immigrant children; determine how the health profile of first- and second-generation international immigrant children has changed between Censuses and establish whether these patterns vary according to children’s age, sex and family socio-economic status.

## Methods

### Data sources

We used the Office for National Statistics Longitudinal Study^21^ (LS) Census records from 1991, 2001 and 2011 linked to birth records. The LS is a dynamic, linked dataset of approximately 1% of the population of England and Wales. The LS links data from each decennial Census, starting in 1971, to birth, cancer registration and mortality registration data for each individual who has been sampled for inclusion. The latest Census data linked to the LS is from 2011.

### Study population and follow-up

This study included all LS members, born between 1973 and 2011, regularly present in a household, and aged <18 years old at the Census date at any of the 1991, 2001 or 2011 Censuses.

### Outcome measures

The Census for England and Wales contains data on two health indicators which are either caregiver-reported or self-reported: limiting long term illness (1991, 2001 and 2011) and general health (reported in 2001 and 2011; see box 1).

**Box 1: Definitions of outcome measures**

<u>Presence of limiting long-term illness</u>

Identified using the following Census questions:

- 1991: *‘Do you have any long-term illness, health problem or handicap which limits your daily activities or the work you can do? Include problems that are due to old age’’* Yes/no
- 2001: *‘Do you have any long-term illness, health problem or disability which limits your daily activities or the work you can do? Include problems that are due to old age’* Yes/no
- 2011: ‘*Are your day-to-day activities limited because of a health problem or disability which has lasted, or is expected to last, at least 12 months? Including problems related to old age’* Yes, limited a lot/Yes, limited a little/No (2011)

<u>Self/caregiver-reported general health</u>

Identified using the following Census questions:

- 1991: not asked
- 2001: *‘Over the last twelve months would you say your health on the whole has been: Good? Fairly Good? Not Good?’* [invited to tick one option]
- 2011: *‘How is your health in general?’* Very good/Good/Fair/Bad/Very bad

We derived a binary variable indicating presence of long-term illness (yes/no) across all three censuses. For general health, the response categories were not directly comparable between 2001 and 2011. Despite this, we derived a binary variable across the two Censuses: poor health (yes/no). The ‘poor health’ category included fairly good and not good in 2001 and fair, bad and very bad in 2011, similar to a previous study.^22^

Note that there is no guidance on the age at which children should complete the Census form or if and from what age their parents/carers need to consult with them before answering the questions about health. The UK Census used a concept of a ‘head of household’ in 1991 and a ‘household reference person’ from 2001 onwards. It is likely that the head of household filled out the census forms for all members of the household. In cases where children in the household have better knowledge of English than for parents/carers, children may fill out the Census form.

### Primary exposure: children’s immigration history

There is no universally agreed definition of an international immigrant, and international immigrants are defined differently across Office for National Statistics datasets.^23^ In this study, we used children’s and their parents’ country of birth (irrespective of nationality or passports held) to define our exposure variable, indicating a history of first or second generation immigration. Children’s country of birth is recorded on the Census form, and parents’ countries of birth are recorded at birth registration. We derived a five-category primary exposure variable; child born

i. in the UK, both parents born in the UK (non-immigrant)
ii. in the UK, mother born outside the UK, father in UK (second generation immigrant via mother only)
iii. in the UK, father born outside the UK, mother in the UK (second generation immigrant via father only)
iv. in the UK, both parents born outside the UK (second generation immigrant via both parents)
v. outside the UK (1^st^ generation immigrant)

UK-born children who were solely registered (that is, with no second parent on the birth certificate) were classified according to their mother’s country of birth. A small proportion of children who were born in the UK according to their Census record did not have a linked birth record, likely due to linkage error. We report these proportions but excluded this group from further analyses.

### Effect modifiers

We examined age group, sex and socio-economic status as the effect modifiers in this study due to the relatively small number of children reporting each outcome.

Child’s age on each respective census day was grouped into a four-category variable, <5, 5-9, 10-14, 15-17 years. Respondents were asked ‘What is your sex?’ in 2001 and 2011, with ‘male’ and ‘female’ as response options, and asked to tick a box for male or female in 1991. We used household tenure to indicate socio-economic status. Household tenure is a well-recognised indicator of socio-economic status in the UK,^24^ and it is the most consistently recorded socio-economic status indicator across the three censuses.^25^ We grouped household tenure into the following four-category variable: owned (owns outright, owns with mortgage); privately rented (via estate agent, employer, friends, family or other rented); socially rented (via local authority, housing association), lives rent free (only available in 2001 and 2011 Census). Communal establishments were coded as such in the 1991 Census but were coded as missing in 2001 and 2011, as a separate Census form was used. We therefore coded the small number of children living in communal establishments in 1991 as having a missing tenure for consistency.

### Statistical analyses

Analyses were carried out separately for each Census. For children in each Census, we described the most common countries of birth and key cohort characteristics.

We calculated the proportion of children who reported long-term illness or poor health according to immigration history. We examined whether any differences were statistically significant using χ*2* tests; *p*<0.05 was deemed to be statistically significant. We fitted logistic regression models, first with no covariates (null models) for each health measure separately. We then included immigration history in the models and examined whether this significantly improved model fit. If immigration history improved the fit of a particular model, we presented these results, and further included effect modification terms for age, sex and household tenure (in separate models). We used Likelihood Ratio tests to determine whether model fit significantly improved; again, with *p*<0.05 deemed to be statistically significant. Model results are presented using odds ratios compared to the grand mean. We used complete case analyses for all models due to the small number of children with missing data. All analyses were carried out in Stata version 17.^26^

## Data Availability

All data used in this study can be requested from the Centre for Longitudinal Study Information and User Support (CeLSIUS): https://www.ucl.ac.uk/epidemiology-health-care/research/epidemiology-and-public-health/research/health-and-social-surveys-research-group/studies-9

## Ethical considerations

This project was approved by the Office for National Statistics Research Accreditation Panel, reference number 2002967.

## Results

Following initial exclusions, the study sample included 121,109, 118,318 and 117,116 children in the 1991, 2001 and 2011 Census, respectively (Figure S1).

The percentage of children who were first-or second-generation immigrants increased from 16.6% (20120 of 121,109 children) in the 1991 Census to 24.7% in the 2011 Census (28965 of 117116 children; Table S1). By 2011, 6912 children (5.9%) were first generation immigrants; and 22053 children (18.8%) were second generation immigrants. Around half (47.4% of 22053) of second-generation immigrant children in 2011 had two parents born outside the UK. The most common country of birth among non-UK born children in each Census was Germany in 1991-2001 and Poland in 2011 (Appendix Table S2). In 1991, the most common countries of birth for mothers and fathers were India and Bangladesh respectively, and in 2001 and 2011, Pakistan for both mothers and fathers.

### Long-term illness

The proportion of children with long-term illness was 2.3%, 4.2% and 3.9% in the 1991, 2001 and 2011 Census respectively (relating to 2696, 4992 and 4524 children in each Census; Table S1). There was no statistically significant difference in the proportion of children with long-term illness and immigration status before 2011 (Table 1).

**Table 1.**
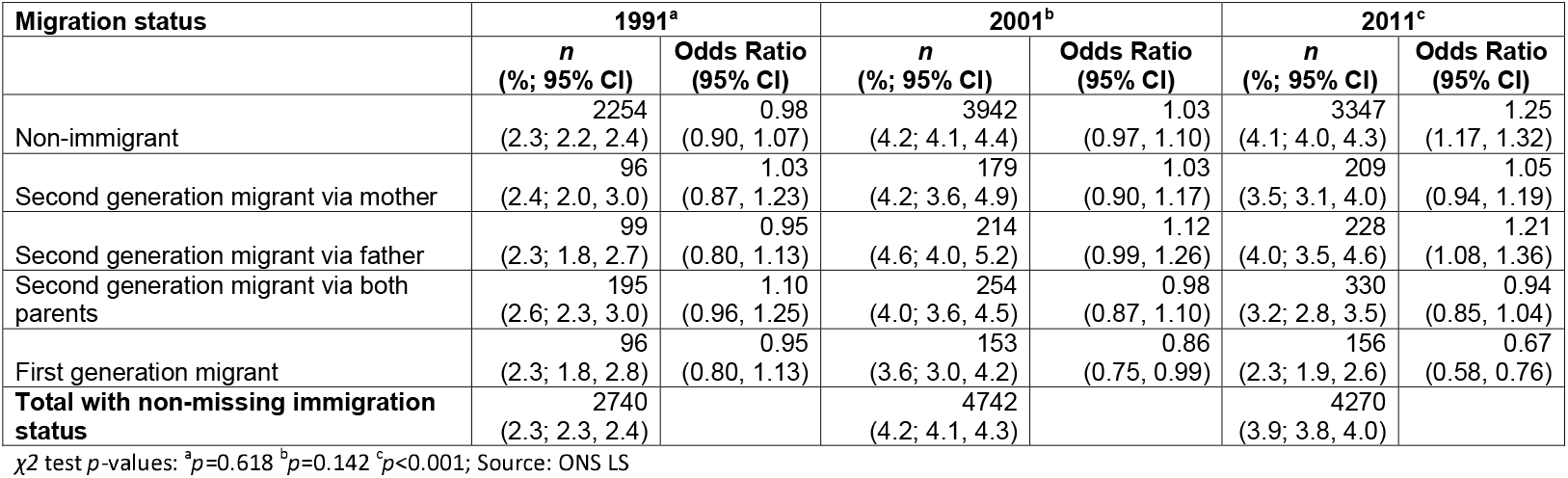
Number and percentages (with 95% confidence intervals) of children with long-term limiting illness and ORs for long-term illness compared to grand mean, according to immigration history and Census year.

| Migration status | 1991<sup>a</sup> |  | 2001<sup>b</sup> |  | 2011<sup>c</sup> |  |
| --- | --- | --- | --- | --- | --- | --- |
|  | *n*<br>(%; 95% CI) | Odds Ratio<br>(95% CI) | *n*<br>(%; 95% CI) | Odds Ratio<br>(95% CI) | *n*<br>(%; 95% CI) | Odds Ratio<br>(95% CI) |
| Non-immigrant | 2254<br>(2.3; 2.2, 2.4) | 0.98<br>(0.90, 1.07) | 3942<br>(4.2; 4.1, 4.4) | 1.03<br>(0.97, 1.10) | 3347<br>(4.1; 4.0, 4.3) | 1.25<br>(1.17, 1.32) |
| Second generation migrant via mother | 96<br>(2.4; 2.0, 3.0) | 1.03<br>(0.87, 1.23) | 179<br>(4.2; 3.6, 4.9) | 1.03<br>(0.90, 1.17) | 209<br>(3.5; 3.1, 4.0) | 1.05<br>(0.94, 1.19) |
| Second generation migrant via father | 99<br>(2.3; 1.8, 2.7) | 0.95<br>(0.80, 1.13) | 214<br>(4.6; 4.0, 5.2) | 1.12<br>(0.99, 1.26) | 228<br>(4.0; 3.5, 4.6) | 1.21<br>(1.08, 1.36) |
| Second generation migrant via both parents | 195<br>(2.6; 2.3, 3.0) | 1.10<br>(0.96, 1.25) | 254<br>(4.0; 3.6, 4.5) | 0.98<br>(0.87, 1.10) | 330<br>(3.2; 2.8, 3.5) | 0.94<br>(0.85, 1.04) |
| First generation migrant | 96<br>(2.3; 1.8, 2.8) | 0.95<br>(0.80, 1.13) | 153<br>(3.6; 3.0, 4.2) | 0.86<br>(0.75, 0.99) | 156<br>(2.3; 1.9, 2.6) | 0.67<br>(0.58, 0.76) |
| **Total with non-missing immigration status** | 2740<br>(2.3; 2.3, 2.4) |  | 4742<br>(4.2; 4.1, 4.3) |  | 4270<br>(3.9; 3.8, 4.0) |  |

$\chi^2$ test *p*-values: <sup>a</sup>*p*=0.618 <sup>b</sup>*p*=0.142 <sup>c</sup>*p*<0.001; Source: ONS LS

In 2011, first generation immigrant children had the lowest prevalence of long-term illness (2.3%; table 1) and non-immigrant children and children who were first generation migrants via their father had the highest prevalence (4.1% and 4.0% respectively). These proportions were significantly different to the grand mean. Inclusion of immigration history significantly improved the fit of the model for long-term illness (LR-test *p*<0.001).

Inclusion of effect modification terms for sex, age group and housing tenure in 2011 significantly improved model fit for long-term illness (LR-test p<0.001 for all three models). Children aged <5 years old, girls and children in families that owned their homes outright were least likely to report long-term illness (Figure 1), irrespective of immigration status.

**Figure 1.**
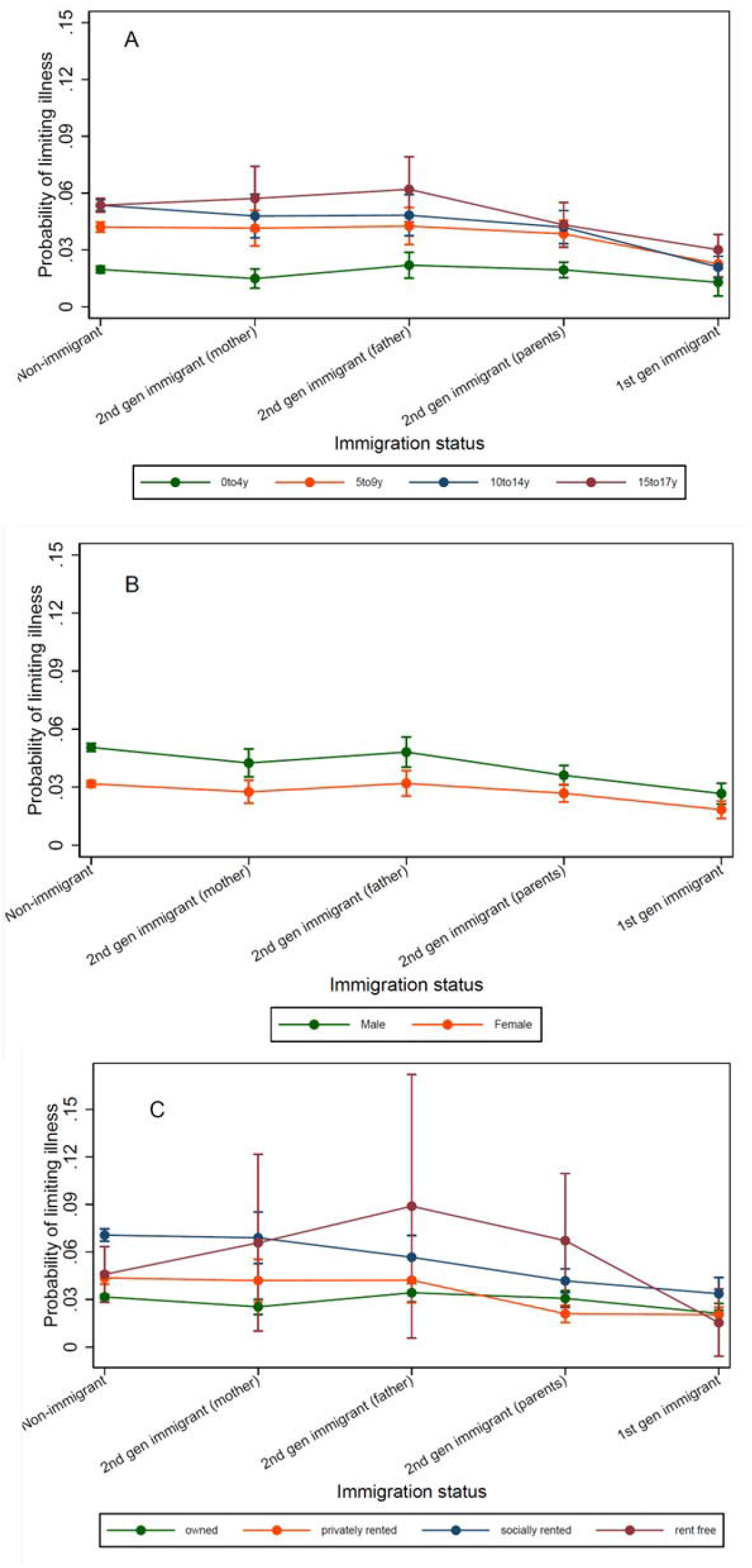
Predicted probabilities of limiting long-term illness by immigration history and age group (A), sex (B) and household tenure, 2011; Source: ONS LS.

### Poor health

The proportion of children with poor health was 9.3% in 2001 and 2.8% in 2011 (relating to 10650 children and 3111 children respectively)

In 2001, first generation immigrant children and non-immigrant children had the lowest reported prevalence of poor health (9.3% and 9.2% respectively; Table 2), whereas children second generation immigrant children via both parents had the highest (12.3%). These differences were significantly different to the grand mean, and inclusion of immigration history significantly improved the fit of the model for poor health (LR-test *p*<0.001).

**Table 2.** Number and percentages (with 95% confidence intervals) of children with poor health and ORs for poor health compared to grand mean, according to immigration history and Census year.

| Poor health |  |  |  |  |
| --- | --- | --- | --- | --- |
| Migration status | 2001<sup>d</sup> |  | 2011<sup>e</sup> |  |
|  | *n*<br>(%; 95% CI) | Odds Ratio<br>(95% CI) | *n*<br>(%; 95% CI) | Odds Ratio<br>(95% CI) |
| Non-immigrant | 8608<br>(9.3; 10.1, 10.5) | 0.92<br>(0.88, 0.96) | 2213<br>(2.7; 2.6, 2.8) | 0.92<br>(0.86, 0.98) |
| Second generation migrant via mother | 395<br>(9.3; 8.5, 10.2) | 0.92<br>(0.85, 1.01) | 169<br>(2.9; 2.4, 3.3) | 0.96<br>(0.84, 1.10) |
| Second generation migrant via father | 480<br>(10.3; 9.4, 11.2) | 1.03<br>(0.95, 1.12) | 179<br>(3.2; 2.7, 3.7) | 1.07<br>(0.94, 1.22) |
| Second generation migrant via both parents | 773<br>(12.3; 11.5, 13.1) | 1.26<br>(1.17, 1.35) | 369<br>(3.5; 3.2, 3.9) | 1.20<br>(1.09, 1.32) |
| First generation migrant | 394<br>(9.2; 8.3, 10.1) | 0.91<br>(0.83, 0.99) | 181<br>(2.6; 2.3, 3.0) | 0.88<br>(0.78, 1.00) |
| **Total with non-missing immigration status** | 10650<br>(9.5; 9.3, 9.6) |  | 3111<br>(2.8; 2.7, 2.9) |  |

$\chi^2$ test $p$ -values: <sup>d</sup> $p < 0.001$ <sup>e</sup> $p < 0.001$ ; Source: ONS LS

We identified statistically significant effect modification by age group and tenure category (Figure 2; LR-test<*p*.0.001 for both models). Children aged 15-17 years old had the highest prevalence of poor health across all immigration histories. As for long-term illness, children living in households that privately own their homes had the lowest prevalence of poor health across all immigration backgrounds.

**Figure 2.**
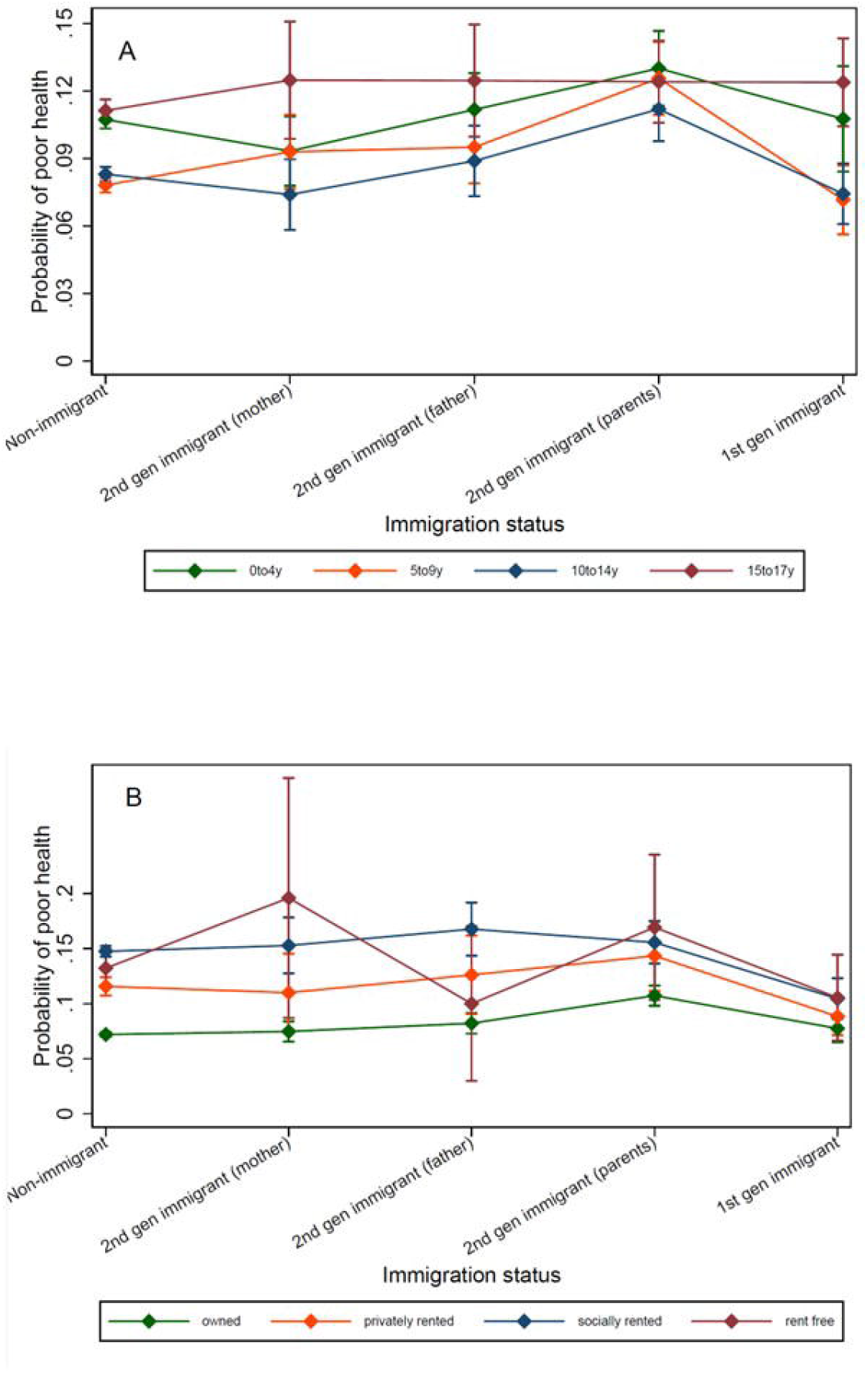
Predicted probabilities of long-term illness health by immigration history and age group (A) and household tenure (B), 2001. Note different y-axis for household tenure; Source: ONS LS.

In 2011, first generation immigrant children and non-immigrant children had the lowest prevalence of poor health (2.6% and 2.7% respectively), and children whose parents were born abroad had the highest prevalence (3.5%). These proportions were significantly different to the grand mean. Inclusion of immigration history significantly improved the fit of the model for poor health (LR-test *p*<0.001). We identified significant effect modification by age group, sex and tenure category respectively for the association between immigration history and poor health.

Predicted prevalence of poor health was highest in children aged 0-4 years among first generation immigrant children, but in 15-17-year-olds among non-immigrant children (Figure 3). Patterns of predicted probabilities of poor health by migration status, sex and household tenure were similar to those for long-term illness.

**Figure 3.**
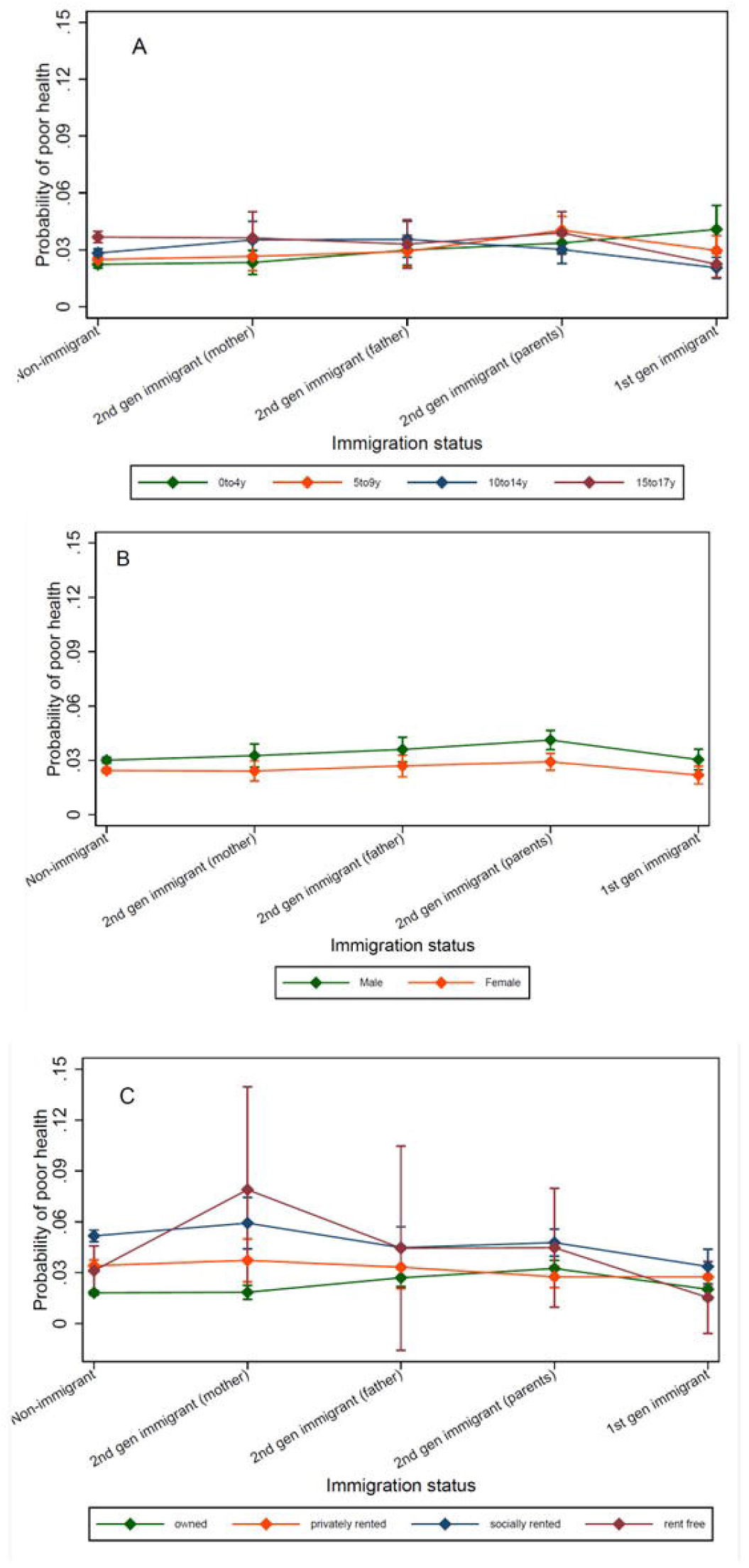
Predicted probabilities of poor health by immigration history and age group, sex and household tenure, 2011; Source: ONS LS.

## Discussion

We found no difference in the prevalence of long-term illness before 2011 according to children’s immigration history. In the 2011 Census, although less than 5% of children overall reported long-term illness, prevalence was highest among non-immigrant children and second-generation immigrant children via their father, and lowest among first-generation immigrant children. In 2001 and 2011, children born in the UK who had two parents born abroad had the highest prevalence of poor health, whereas prevalence was lowest among non-immigrant and first-generation immigrant children. In 2001, children aged 15-17 years had the highest prevalence of poor health irrespective of immigration history. In 2011, poor health prevalence was highest for 15-17-year-old non-immigrant children but in the pre-school age group among first-generation immigrant children.

To our knowledge, this is the largest study to date in the UK, and one of the largest studies internationally, of self-or caregiver-reported health in children according to immigration history. The longevity of data availability within the LS allowed us to explore trends over a 20-year period since 1991. Linkage between birth records and Census data supported comparison of health outcomes between first- and second-generation immigrant children, since country of birth of parents is not asked about in the Census. Similar studies have been possible in Scandinavia via multi-generational population registers,^27 28^ which do not exist in the UK. The LS contains data for 1% of the population of England and Wales. This was sufficiently large to allow us to detect differences in two child health outcomes according to immigration history. However, given the relatively low proportion of children with either long-term illness or poor health, national, linked Census and birth registration data are required to establish differences according to specific regions or countries of origin, and to take into account ethnicity and length of stay in the UK.^29^ Further, linkage to primary and secondary care data would allow researchers to compare self-or caregiver reported health with use of health services. These analyses may become possible using the Public Health Data Asset developed by the Office for National Statistics,^30^ once linked health data are made available to external researchers.

We only had access to LS data until 2011, as the LS is currently being updated with 2021 Census data. We also did not link children’s records over time to examine evolving health trajectories according to children’s immigration history. Once 2021 Census data are available, our analyses should be updated to examine whether our findings have changed, and how children’s health shift as they grow up. Although it is a legal requirement to take part in the UK Census, immigrants, and particularly newly arrived immigrants, may be less likely to respond.^31^ Further, health-related questions may be interpreted differently according to immigration history due different cultural beliefs or views on health.^32^ The wording of questions has also changed between Censuses. Specifically, the question regarding health status used response categories that were not strictly comparable over time.

Only a small proportion of children had limiting long-term illness. However, we found that differences in long-term illness according to immigration history became more prominent over time. This may be due to changes in the most common sending countries for parents and children, changes in migration policy, or changes in the underlying prevalence of long-term illness, due to increasing survival of children with disability,^33^ improved detection and diagnosis (for example for neurodiversity)^34^ or an increase in the prevalence of mental health conditions.^35^

By 2011, we identified lower prevalence of long-term illness among first generation immigrant children compared to children who are non-immigrants, however the prevalence of poor health was similar in these groups in both 2001 and 2011. These results would generally support a healthy migrant effect for self/caregiver-reported health status for children in the UK. Our findings are therefore in line with previous research showing a similar effect for self-reported health among adults in the UK, with the largest differences being seen in the prevalence of long-term illness.^11^ Our results contradict those of a large, global systematic review which found that immigrant children had generally worse health than children in the host population, for outcomes including malnutrition, mental health and mortality. However, relative health outcomes for immigrants vary according to both sending and host setting, outcomes considered and time since migration, which may explain our differential findings.

Second generation immigrant children whose father was born outside the UK had similar prevalence of long-term illness to non-immigrant children, whereas children whose parents were both born outside the UK were more likely to report poor health than non-immigrant children. These results contrast with our previous study showing lower risk of emergency hospital admissions in preschool children whose parents were born abroad.^8^ However, they concur with previous studies showing increasing prevalence of chronic illness and a reduction of the healthy migrant effect in children with successive generations since migration, sometimes referred to as assimilation.^36^ A Finnish study showed that health service contacts were higher for children whose fathers, rather than mothers, were born abroad, particularly for mental health conditions. ^37^ A Danish study similarly showed that children with one parent born abroad were more likely to require mental health-related secondary care contacts than non-immigrant or first generation immigrant children.^38^ The England and Wales Census does not allow respondents to report the type of illnesses experienced. Linking LS to primary and secondary care data would therefore allow exploration into the type of health conditions where observed disparities according to immigration history are greatest.

First generation-immigrant children had lower prevalence of long-term illness but similar prevalence of poor health as non-immigrant children. Children whose father, or both parents, were born outside the UK were more likely to have long-term illness and poor health respectively. The UK Health Security Agency publishes advice to health professionals supporting adults^20^ and children^39^ who have moved to the UK. Future research into which particular health conditions account for the increased risk of poor health and long-term illness for second generation immigrant children could inform clinical guidelines to support this group accessing and using the National Health Service.

## Acknowledgements

*The permission of the Office for National Statistics to use the Longitudinal Study is gratefully acknowledged, as is the help provided by staff of the Centre for Longitudinal Study Information × User Support (CeLSIUS). CeLSIUS is funded by the ESRC under project ES/V003488/1. The authors alone are responsible for the interpretation of the data*.

*This work contains statistical data from ONS which is Crown Copyright. The use of the ONS statistical data in this work does not imply the endorsement of the ONS in relation to the interpretation or analysis of the statistical data. This work uses research datasets which may not exactly reproduce National Statistics aggregates*.

