## Supplementary Material for "Health of first- and second-generation immigrant children in England and Wales: serial, cross-sectional study using linked Census and birth registration data"

*Table S1: Distribution of key variables among children in each Census: Source: ONS LS*

|  | **Census** | | |
| --- | --- | --- | --- |
| **Characteristic** | **1991** | **2001** | **2011** |
| **Migration status** | **n (%)** | | |
| Non-immigrant | 96943 (80.1) | 92991 (78.6) | 81016 (69.2) |
| Second generation migrant via mother | 3936 (3.3) | 4241 (3.6) | 5935 (5.1) |
| Second generation migrant via father | 4406 (3.6) | 4671 (4) | 5668 (4.8) |
| Second generation migrant via both parents | 7523 (6.2) | 6312 (5.3) | 10450 (8.9) |
| First generation migrant | 4255 (3.5) | 4302 (3.6) | 6912 (5.9) |
| Born in UK, birth record missing | 4046 (3.3) | 3700 (3.1) | 4118 (3.5) |
| Country of birth (child) missing | 0 (0) | 2101 (1.8) | 3017 (2.6) |
| **Age group** |  |  |  |
| 0 to 4 years | 35680 (29.5) | 30024 (25.4) | 33762 (28.8) |
| 5 to 9 years | 33591 (27.7) | 32997 (27.9) | 31252 (26.7) |
| 10 to 14 years | 31939 (26.4) | 34860 (29.5) | 31759 (27.1) |
| 15 to 17 years | 19899 (16.4) | 20422 (17.3) | 20343 (17.4) |
| Missing | 0 (0) | 15 (0) | 0 (0) |
| **Sex** |  |  |  |
| Male | 61778 (51) | 60250 (50.9) | 59657 (50.9) |
| Female | 59331 (49) | 58068 (49.1) | 57459 (49.1) |
| **Household tenure** |  |  |  |
| Owner occupied | 83771 (69.2) | 80953 (68.4) | 71692 (61.2) |
| Privately rented | 7501 (6.2) | 8497 (7.2) | 20180 (17.2) |
| Socially rented | 29341 (24.2) | 26327 (22.3) | 23805 (20.3) |
| Lives rent free* |  | 2076 (1.8) | 1007 (0.9) |
| Missing | 496 (0.4) | 465 (0.4) | 432 (0.4) |
| **Presence of limiting long-term illness** |  |  |  |
| No | 118260 (97.7) | 113037 (95.5) | 111994 (95.6) |
| Yes | 2849 (2.4) | 4992 (4.2) | 4524 (3.9) |
| Missing | 0 (0) | 289 (0.2) | 598 (0.5) |
| **General health**** |  |  |  |
| Good |  | 106963 (90.4) | 113170 (96.6) |
| Poor |  | 11070 (9.4) | 3348 (2.9) |
| Missing |  | 285 (0.2) | 598 (0.5) |
| **Total** | 121109 (100) | 118318 (100) | 117116 (100) |

*General health was not collected in 1991

*Table S2. Most common countries of birth of non-UK born children, mothers and fathers (if stated), by Census (n indicates number of children): Source: ONS LS*

| **1991** | | | | | |
| --- | --- | --- | --- | --- | --- |
| **Children** | | **Mothers** | | **Fathers** | |
| **Country** | ***n*** | **Country** | ***n*** | **Country** | ***n*** |
| Germany | 555 | India | 1,931 | Bangladesh | 2,103 |
| Bangladesh | 521 | Pakistan | 1,737 | Pakistan | 1,801 |
| Pakistan | 372 | Irish Republic | 1,202 | Irish Republic | 1,211 |
| USA | 316 | Kenya | 533 | Cyprus | 805 |
| Irish republic | 238 | Jamaica | 491 | Barbados | 593 |
| Australia | 147 | Bangladesh | 481 | Zimbabwe | 592 |
| South Africa | 141 | Germany | 455 | Germany | 323 |
| India | 118 | Cyprus | 272 | Italy/San Marino | 274 |
| Africa - rest | 112 | Hong Kong | 247 | Malasia/Singapore | 235 |
| Hong Kong | 99 | Malasia/Singapore | 242 | Tanzania | 234 |
| **2001** | | | | | |
| **Children** | | **Mothers** | | **Fathers** | |
| **Country** | ***n*** | **Country** | ***n*** | **Country** | ***n*** |
| Germany | 423 | Pakistan | 1797 | Pakistan | 2,088 |
| U.S.A | 307 | India | 1304 | Bangladesh | 1321 |
| Pakistan | 242 | Bangladesh | 785 | Irish Republic | 644 |
| South Africa | 220 | Irish Republic | 612 | India | 605 |
| Somalia | 184 | Germany | 601 | Cyprus | 520 |
| Bangladesh | 178 | Kenya | 460 | Germany | 431 |
| Hong Kong | 132 | Malasia/Singapore | 280 | Zimbabwe | 372 |
| India | 131 | Jamaica | 259 | Kenya | 296 |
| Republic of Ireland | 124 | Nigeria | 241 | Malasia/Singapore | 274 |
| France | 113 | Uganda | 213 | Barbados | 234 |
| **2011** | | | | | |
| **Children** | | **Mothers** | | **Fathers** | |
| **Country** | ***n*** | **Country** | ***n*** | **Country** | ***n*** |
| Poland | 715 | Pakistan | 2,291 | Pakistan | 2,744 |
| India | 418 | India | 1416 | Bangladesh | 1,399 |
| United States | 350 | Bangladesh | 1,290 | India | 1303 |
| Pakistan | 344 | Poland | 775 | Nigeria | 614 |
| Germany | 290 | Germany | 696 | Poland | 558 |
| Somalia | 289 | Somalia | 600 | Germany | 551 |
| France | 188 | Nigeria | 521 | Somalia | 539 |
| South Africa | 181 | Republic of Ireland | 462 | Republic of Ireland | 432 |
| Nigeria | 180 | Sri Lanka | 378 | Jamaica | 430 |
| Netherlands | 179 | South Africa | 370 | Sri Lanka | 407 |

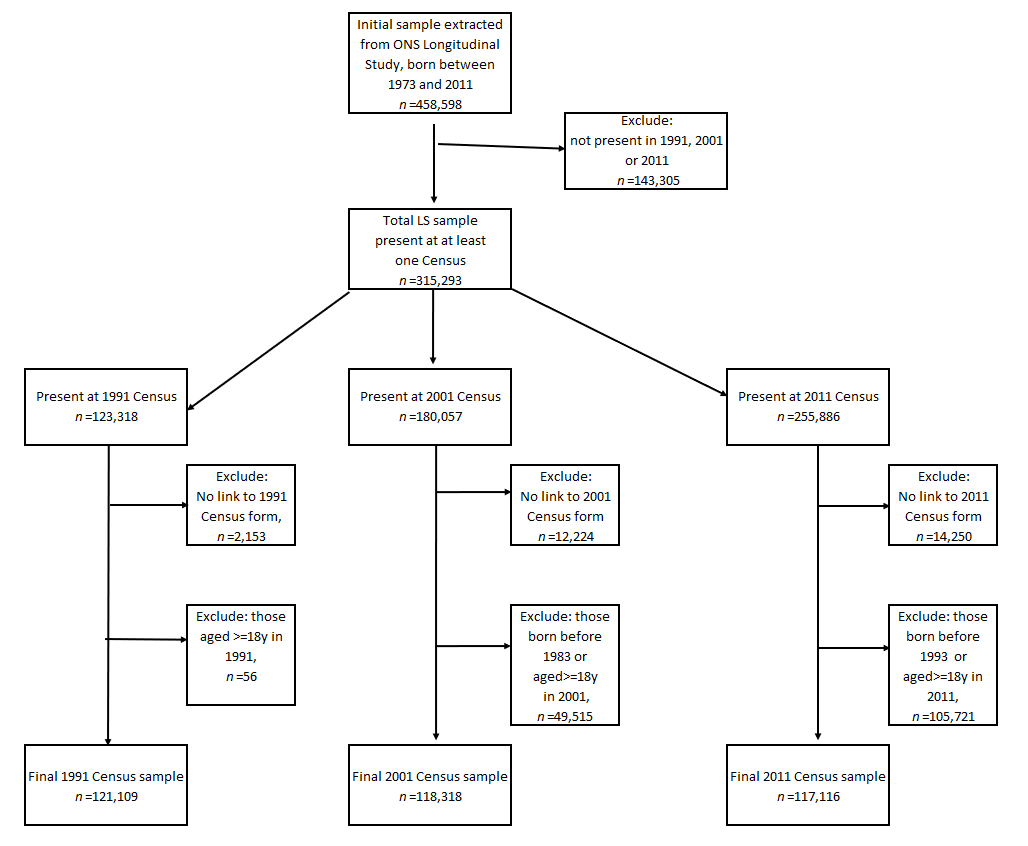
 *Figure S1. Flowchart of derivation of final study sample: Source: ONS LS*
